# Electrophysiological connectivity markers of preserved language functions in post-stroke aphasia

**DOI:** 10.1101/2021.11.23.21266546

**Authors:** Priyanka Shah-Basak, Gayatri Sivaratnam, Selina Teti, Tiffany Deschamps, Aneta Kielar, Regina Jokel, Jed A. Meltzer

**Author notes:** **Corresponding author: Priyanka P. Shah-Basak, PhD**, Department of Neurology, Medical College of Wisconsin, 8701 W. Watertown Plank Rd, Wauwatosa, WI 53226 USA. Note that the current address of the first author (Shah-Basak) is Department of Neurology, Medical College of Wisconsin, Milwaukee, WI 53226 USA.

## Abstract

Post-stroke aphasia is a consequence of localized stroke-related damage as well as global disturbances in a highly interactive and bilaterally-distributed language network. Aphasia is increasingly accepted as a network disorder and it should be treated as such when examining the reorganization and recovery mechanisms after stroke. In the current study, we sought to investigate reorganized patterns of electrophysiological connectivity, derived from resting-state magnetoencephalography (rsMEG), in post-stroke chronic (>6 months after onset) aphasia. We implemented amplitude envelope correlations (AEC), a metric of connectivity commonly used to describe slower aspects of interregional communication in resting-state electrophysiological data. The main focus was on identifying the oscillatory frequency bands and frequency-specific spatial topology of connections associated with preserved language abilities after stroke.

RsMEG was recorded for 5 minutes in 21 chronic stroke survivors with aphasia and in 20 matched healthy controls. Source-level MEG activity was reconstructed and summarized within 72 atlas-defined brain regions (or nodes). A 72×72 leakage-corrected connectivity (of AEC) matrix was obtained for frequencies from theta to gamma (4–128 Hz). Connectivity was compared between groups, and, the correlations between connectivity and subscale scores from the Western Aphasia Battery (WAB) were evaluated in the stroke group, using partial least squares analyses. Posthoc multiple regression analyses were also conducted on a graph theory measure of node strengths, derived from significant connectivity results, to control for node-wise properties (local spectral power and lesion sizes) and demographic and stroke-related variables.

Connectivity among the left hemisphere regions, i.e. those ipsilateral to the stroke lesion, was greatly reduced in stroke survivors with aphasia compared to matched healthy controls in the alpha (8-13 Hz; p=0.011) and beta (15-30 Hz; p=0.001) bands. The spatial topology of hypoconnectivity in the alpha vs. beta bands was distinct, revealing a greater involvement of ventral frontal, temporal and parietal areas in alpha, and dorsal frontal and parietal areas in beta. The node strengths from alpha and beta group differences remained significant after controlling for nodal spectral power. AEC correlations with WAB subscales of object naming and fluency were significant. Greater alpha connectivity was associated with better naming performance (p=0.045), and greater connectivity in both the alpha (p=0.033) and beta (p=0.007) bands was associated with better speech fluency performance. The spatial topology was distinct between these frequency bands. The node strengths remained significant after controlling for age, time post stroke onset, nodal spectral power and nodal lesion sizes.

Our findings provide important insights into the electrophysiological connectivity profiles (frequency and spatial topology) potentially underpinning preserved language abilities in stroke survivors with aphasia.

## 1. INTRODUCTION

Stroke recovery is thought to depend on neuroplastic changes within preserved brain areas (Saur et al., 2006). In aphasia, language recovery relies on spared left-hemispheric areas surrounding the stroke lesion (perilesional), and/or right-hemispheric contralesional areas (Fridriksson et al., 2010; Chrysikou and Hamilton, 2011; Hamilton et al., 2011; Turkeltaub et al., 2011). Evidence also suggests that the functional capacity of perilesional areas is compromised due to the adjacent lesion (Tecchio et al., 2006; Meinzer et al., 2008). Using resting-state magnetoencephalography (rsMEG) in patients with aphasia, we and others have shown that spontaneous neural activity in perilesional areas is abnormal, exhibiting shifts toward slower oscillatory activity and alterations in signal complexity (Laaksonen et al., 2013; Chu et al., 2015; Kielar et al., 2016; Shah-Basak et al., 2020). Physiological slow activity manifests as increased delta (1-4 Hz) and theta (4-7 Hz) spectral power but reduced beta (15-30 Hz) and low-gamma (30-50 Hz) power. It is hypothesized that such slowing indicates dysfunction in otherwise intact brain tissue, which may hinder recovery processes.

To date, much work has focused on local dysfunction by analyzing changes in regional spectral power and signal complexity after stroke. But little is known about electrophysiological abnormalities manifesting in interregional connectivity, encompassing the perilesional and contralesional areas in post-stroke aphasia. We know that processing of language relies on dynamic and rapid processes emerging across a large network of brain regions (Piai and Zheng, 2019). Coherent neural oscillations via phase synchronization or correlated oscillatory amplitude (or power envelope) fluctuations are thought to constitute the physiological mechanism that supports timely coordination, signaling and communication across distant brain regions (Fries, 2005; Wang, 2010; Fries, 2015; Cox et al., 2018). In the current study, we sought to characterize spontaneous (resting-state) long-range oscillatory connectivity using a metric of amplitude envelope correlation (AEC) in frequency bands ranging from theta to low-gamma (4-50 Hz), and evaluated relationships between connectivity and language outcomes in stroke survivors with aphasia. Amplitude-based connectivity during the resting state is a widely studied phenomenon, wherein oscillatory amplitude or power envelope fluctuations are thought to capture the slower aspects of interregional communication (Cox et al., 2018).

Recent work in aphasia underscores functional network-level impairments after stroke (Grefkes and Fink, 2011; Carrera and Tononi, 2014). Stroke induces disruptions in the lesion zone and in functionally connected areas far from the lesion that may be critical for language (Turken and Dronkers, 2011). Disconnections can contribute to specific linguistic impairments, due to disruptions in relevant information transfer across brain regions in language subnetworks. Plasticity after stroke is associated with synchronization of spontaneous neural oscillations between brain areas. One study found that greater oscillatory synchronization of language and motor areas with the rest of the cortex at 2-3 weeks after stroke is linked to improvement in corresponding clinical functions during subsequent weeks (Nicolo et al., 2015). In particular, coherent synchrony in alpha and beta frequencies has been associated with post-stroke cognitive and motor recovery (Westlake et al., 2012; Dubovik et al., 2013; Petrovic et al., 2017). This is an emerging field of research but so far very little is known about the long-range connectivity patterns underpinning spared language functions in post-stroke aphasia.

In this study, we implemented rsMEG analysis for computing AEC to characterize electrophysiological connectivity differences between 21 stroke patients with chronic aphasia and 20 age- and sex-matched healthy controls. Based on prior literature, we expected *hypo*connectivity in the alpha (8-13 Hz) and beta (15-30 Hz) bands in the left (or ipsilesional) hemisphere in patients compared to the healthy controls. For the group differences, there was little doubt that we would find connectivity differences emerging from the lesioned hemisphere. Thus, the primary interest of this analysis was to determine the involvement of specific frequency bands, and the spatial topology of connections (reflecting the number of connections, and edges or links between brain areas in a network) within those frequency bands. The second aim was to disentangle the relationship between frequency-specific connectivity patterns and preserved language functions after stroke. We hypothesized that greater connectivity in the alpha and beta bands and connections involving the left perilesional areas would positively correlate with language performance. For these analyses, we compared AEC estimates with subscales of a widely used clinical language battery—the Western Aphasia Battery (WAB)—including selected measures of fluency, auditory comprehension, repetition and object naming (Kertesz, 2007). Finally, to control for potential confounding effects of stroke-related variables, post-hoc regression analyses were conducted with lesion sizes, time post stroke onset and age as covariates. Overall, our study results provide important insights into the electrophysiological connectivity profiles that underpin preserved language abilities in aphasia. This understanding could inform aphasia treatments, particularly those employing rhythmic brain stimulation to promote language recovery after stroke.

## 2. MATERIALS AND METHODS

### 2.1 Participants

Resting-state MEG data was collected as part of two prior studies (Kielar et al., 2016; Shah-Basak et al., 2020). Twenty-one stroke survivors with chronic aphasia (mean ± standard deviation: age: 61.5±13.2 years; education: 16.3±2.5 years; 16 males; Table 1) and 20 older healthy controls (age: 63.4±12.6 years; education: 17.5±2.4 years; 15 males) were included in the analysis. Patients suffered a single left-hemispheric stroke on average 6 years prior to the study (stroke onset: 6.1±4.7, range: 0.6-21 years). Twelve patients suffered an ischemic stroke, 6 a hemorrhagic stroke, and 3 had an unspecified etiology.

**Table 1.** Demographics, clinical variables and language scores for the stroke patients.

| ID | Age range (y) | Education (y) | Sex | Time post-onset | Etiology | Aphasia type | % left damage | WAB-Fluency | WAB-Comp | WAB-Rep | WAB-Naming |
| --- | --- | --- | --- | --- | --- | --- | --- | --- | --- | --- | --- |
| P1 | 56-60 | 16 | Male | 4y | Ischemic | Non-fluent | 19.8 | 4 | 10 | 7.5 | 10 |
| P2 | 61-65 | 12 | Male | 5y | Ischemic | Fluent | 38.64 | 2 | 10 | 6 | 5.5 |
| P3 | 66-70 | 16 | Male | 6y 2m | Unspecified | Non-fluent | 12.94 | 8 | 10 | 5 | 8.5 |
| P4 | 66-70 | 18 | Male | 9y | Ischemic | Non-fluent | 21.3 | 9 | 10 | 9 | 10 |
| P5 | 71-75 | 19 | Male | 21y 4m | Unspecified | Anomia | 31.81 | 1 | 8 | 1.5 | 1 |
| P6 | 31-35 | 19 | Female | 4y | Hemorrhagic | Conduction | 18.96 | 7 | 9 | 8.5 | 10 |
| P7 | 71-75 | 15 | Female | 9y 4m | Hemorrhagic | Anomia | 23.35 | 8 | 10 | 8 | 9.5 |
| P8 | 61-65 | 12 | Female | 13y | Ischemic | Anomia | 28.71 | 8 | 10 | 7 | 8.5 |
| P9 | 56-60 | 13 | Male | 6y 5m | Hemorrhagic | Non-fluent | 42.1 | 2 | 9 | 5 | 4.5 |
| P10 | 41-45 | 18 | Male | 5y 9m | Ischemic | Non-fluent | 22.29 | 2 | 8 | 5 | 7.5 |
| P11 | 66-70 | 16 | Male | 4y 10m | Unspecified | Conduction | 15.48 | 9 | 9 | 9.5 | 9.5 |
| P12 | 46-50 | 18 | Female |  | Hemorrhagic | Non-fluent | 20.13 | 5 | 9 | 5 | 9 |
| P13 | 46-50 | 18 | Male | 4y 1m | Hemorrhagic | Anomia | 20.87 | 5 | 8 | 7.5 | 9.5 |
| P14 | 71-75 | 15 | Male | 2y 4m | Ischemic | Anomia | 9 | 8 | 10 | 7.5 | 7.5 |
| P15 | 46-50 | 15 | Male | 2y 3m | Ischemic | Conduction | 16.81 | 8 | 9 | 7.5 | 10 |
| P16 | 81-85 | 19 | Male | 10y | Ischemic | Anomia | 1.82 | 9 | 9 | 10 | 9.5 |
| P17 | 76-80 | 20 | Male | 7m | Ischemic | Fluent | 9.81 | 7 | 9 | 6 | 6 |
| P18 | 46-50 | 16 | Male | 4y | Ischemic | Non-fluent | 21.71 | 6 | 10 | 9.5 | 10 |
| P19 | 61-65 | 20 | Male | 6y 1m | Ischemic | Non-fluent | 36.36 | 0 | 7 | 3.5 | 0.5 |
| P20 | 66-70 | 13 | Female | 3y 3m | Hemorrhagic | Anomia | 13.09 | 9 | 9 | 10 | 10 |
| P21 | 66-70 | 15 | Male | 1y | Ischemic | Non-fluent | 23.03 | 4 | 9 | 9 | 9 |
| **Mean (SD)** | **61.5 (13.2)** | **16.3 (2.5)** | **16 Males** | **6y 1m (4y 9m)** |  |  | **21.3 (10.0)** | **5.8 (3.0)** | **9.1 (0.8)** | **7.0 (2.3)** | **7.9 (2.9)** |

y = years; m = months; SD = Standard deviation; WAB = Western Aphasia Battery; WAB-Comp = Auditory Comprehension; WAB-Rep = Repetition; Unspecified - not clear from medical reports

The lesion overlap in stroke survivors, provided in Figure 1, indicated maximum damage (in ∼60% of participants) in the left superior temporal gyrus and rolandic operculum (MNI coordinates: - 54 -28 14 and -40 -18 12). The diagnosis of aphasia was made by a speech-language pathologist and/or a board-certified neurologist, and further verified based on clinical presentation, narrative speech samples and standardized linguistic tests. The healthy controls were matched with the stroke patients on age (p=0.65), education (p=0.13) and sex (75% males).

**Figure 1.**
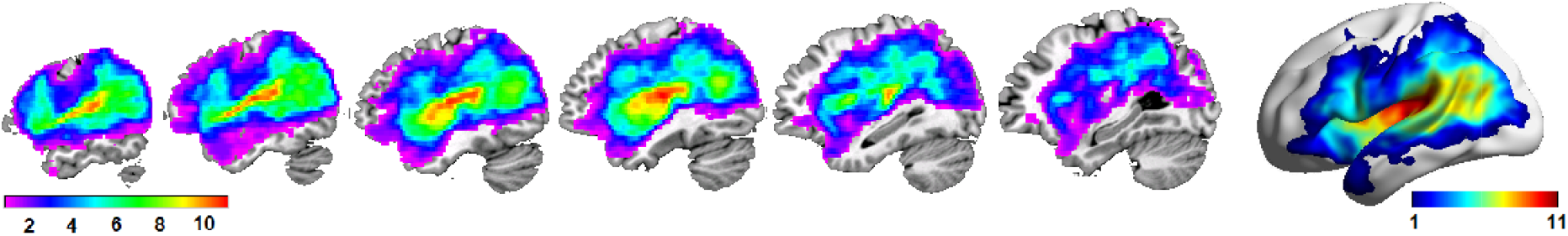
Lesion overlap across 21 stroke survivors with aphasia (thresholded from 1 to 11). Surface representation of overlap is also provided.

All participants were right-handed (pre-stroke), native speakers of English, and had normal hearing and normal or corrected-to-normal vision. All stroke patients retained sufficient language comprehension capacity to consent and follow task instructions. Exclusion criteria were other neurological diseases, language disorders (for controls), head traumas or brain surgery, epilepsy, severe psychiatric disorders, unstable or poor health, and any contraindications to MRI or MEG (Kielar et al., 2016; Kielar et al., 2018).

This study was approved by the Research Ethics Board at our institution. All participants gave their written informed consent according to the Declaration of Helsinki prior to the study and were compensated for their participation.

### 2.2 Clinical language battery

All patients completed the Western Aphasia Battery-Revised (WAB-R) bedside form, including selected subtests of fluency, auditory comprehension, repetition and object naming (Kertesz, 1982, 2007). The WAB fluency test consisted of a description of the ‘Cookie theft’ picture, and a rating based on the sentence length, complexity, speed, and number of paraphasias. WAB comprehension was assessed through yes/no questions, word recognition and sequential commands tasks, and WAB repetition through asking participants to repeat single words, phrases and sentences. WAB object naming consisted of 20 different picture stimuli. The summary scores for each of the subscales are provided in Table 1. In our cohort on average, patients had the least difficulty following auditory commands on the WAB-Comprehension subtest with the scores ranging from 7-10 (mean: 9.1 ± 0.8), but faced most difficulty with the picture description task on the WAB-Fluency subtest with scores ranging from 0-9 (mean: 5.8 ± 3.0).

### 2.3 MRI acquisition

MRI was carried out on a 3-Tesla scanner (Siemens TIM Trio). For MEG source localization, we acquired a T1-weighted MPRAGE image (1mm isotropic voxels, TR = 2000 milliseconds (ms), TE=2.63ms, FOV=256×256mm2, 160 axial slices, scan time 6m, 26s). MR-visible markers were placed at the fiducial points for accurate co-registration with MEG, aided by digital photographs.

### 2.4 MEG acquisition, head modeling and source analysis

Resting-state MEG signals were recorded with a 151-channel whole-head system with axial gradiometers (VSMMedTech, Coquitlam, Canada), acquired continuously for 300 seconds (or 5 minutes) at a sampling rate of 625 Hz or 1250 Hz (down-sampled to 625 Hz) and with an online synthetic 3rd-order gradient noise reduction. Head position with respect to the MEG helmet was monitored using three coils placed at fiducial landmarks of the head (nasion, left and right pre-auricular points). Head positions were measured before and after the resting-state run, and the two positions were averaged. Multi-sphere head models were constructed for MEG source analysis (Huang et al., 1999) using the T1-weighted MRI. Source analysis was conducted on an isotropic grid with a spacing of 10 mm, or a voxel size of 10 mm^3^, across the whole-brain. The Synthetic Aperture Magnetometry (SAM) beamformer (Robinson and Vrba, 1999; Vrba and Robinson, 2001) was implemented using the CTF software (CTF; Port Coquitlam, British Columbia, Canada), and further analyses were supplemented with in-house MATLAB scripts. SAM is a scalar beamformer, in which a nonlinear optimization technique is used to select one direction of current flow at each voxel (or grid location) to maximize dipole power. In short, SAM provides a series of sensor weights for each voxel; the weights are computed so as to pass signal from a dipole located in the target voxel, while minimizing signal power from all other locations. In this study, the beamformer weights were computed at each voxel (10 mm^3^), and were then multiplied with the original sensor time series data to yield a new spatially-filtered signal, the so-called *virtual* time series (Vrba and Robinson, 2001). Normalized weights were computed as a signal-to-noise ratio with noise power estimated as the lowest singular value of the sensor covariance matrix. This step rendered the virtual time series with dimensionless units.

Prior to beamforming, signals were segmented into 4-second epochs, and bandpass filtered from 0.1 to 150 Hz. Raw MEG sensor signals were screened for motion (e.g., coughs, sneezes, yawns, head movements) and environmental artifacts, and epochs containing obvious signal disruptions or drifts were rejected (<1% of all epochs). The SAM beamformer procedure effectively attenuates physiological artifacts such as those from muscle activity and eye movements or blinks (Vrba, 2002; Cheyne et al., 2007). Therefore, the epochs containing these artifacts were not rejected manually.

Voxelwise source signals were then summarized within 72 cortical and subcortical regions (Supplementary Figure 1), as defined by the Automated Anatomical Labeling (AAL) atlas (Tzourio-Mazoyer et al., 2002), using the ‘mean flip’ method implemented in the MNE-Python package (Gramfort et al., 2013), which flips the sign of some voxels prior to averaging to ensure that signals having opposite polarity due to arbitrary selection of the beamformer direction do not cancel each other. Eighteen regions in the AAL atlas were excluded prior to any statistical analysis: (1) due to limited voxelwise coverage (i.e. 0 voxels within an AAL-defined region) based on individual participants’ anatomical scans, which included Heschl’s gyri and amygdala, (2) involving subcortical regions including the basal ganglia regions and the thalamus, and (3) other regions for which we did not have a priori hypotheses in language processes such as the olfactory cortices and the paracentral lobule. In stroke patients, the voxels overlapping with lesion maps in individual patients were excluded, before summarizing the voxelwise data within the AAL-defined regions or nodes.

### 2.5 Amplitude envelope correlations

We used the methods described by Hipp and colleagues (2012) to compute the amplitude envelope correlations or AEC between each of the 72 node pairs. The input signals for this analysis in each participant were the node time series with non-overlapping epochs of 4 seconds. These node signals were downsampled from 625 Hz to 312.5 Hz to reduce the computational demand for subsequent AEC analyses. The downsampled signals were then subjected to spectral analysis using Morlet’s wavelets to obtain signals in the frequency domain. The frequencies of interest (FOI) were logarithmically spaced with base 2 and exponents ranging from 1 to 7 in steps of 0.25 (2 to 128 Hz) (Hipp et al., 2012). The number of cycles for wavelet analysis was set to half of the FOIs. The output of the Morlet’s wavelet analyses was set to single trial complex, resulting in 75 epochs across 72 nodes for 25 FOIs and 1250 time points (4 seconds * 312.5 Hz). To reduce spurious correlations due to signal leakage (O’Neill et al., 2015), the frequency domain signals from each node were orthogonalized to every other node using the ordinary least squares analysis before computing the AECs. The orthogonalization between node pairs was conducted by subtracting the part of the complex signal (say Y) from one node that can be linearly predicted from the complex signal (say X) from another node. As described in detail by Hipp and colleagues (2012), the operation for orthogonalization is as follows:

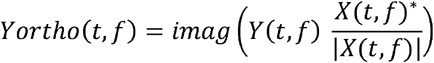

where t are the time points, f is the FOI, * is the complex conjugate, and imag() is the imaginary part of the complex number. Orthogonalization was bi-directional from X to Y and Y to X. The AECs were then computed for both orthogonalized directions using Pearson correlation analysis, and the two resulting correlation coefficients were averaged together for further analyses. At the end of this computation, a 72×72 AEC matrix was obtained for each epoch and FOI. A single AEC matrix per FOI was obtained by averaging across epochs, which was subjected to the Fisher r-to-z transformation. Statistical analyses were conducted on the averages of the AEC matrices within canonical frequency bands—theta (4-7 Hz), alpha (8-13 Hz), beta (15-30 Hz) and low-gamma (25-50 Hz).

### 2.6 Local spectral properties

Power spectral densities of the voxelwise virtual signals were computed using the multitaper method in MATLAB (Thomson, 1982). The time half bandwidth (NW) was set to 3, which resulted in 5 (2*(NW)-1) discrete prolate spheroidal or Slepian sequences for multitaper computations. A frequency resolution of 0.3052 Hz was achieved with FFT length of 1024 and sampling frequency of 625 Hz. Relative power was computed for each canonical frequency band as a ratio of the sum of power within a frequency band and the sum of total power from 1-80 Hz. Relative power estimates were then averaged within the AAL-defined nodes. These estimates were entered as covariates in our statistical analysis to control for the potential contribution of local spectral properties toward connectivity findings.

### 2.7 Statistical Analysis

Partial Least Squares (PLS) (McIntosh et al., 1996) analyses were conducted to compare AEC estimates in each of the frequency bands between stroke patients and age-matched controls. Multivariate methods such as PLS do not necessitate corrections for multiple comparisons because the statistical inference testing is done at the level of a full multivariate pattern rather than at the level of individual nodes (McIntosh and Lobaugh, 2004). For the PLS group comparisons, the input data matrices had *n* rows of participants, nested within groups (*design variables*) and *v* columns of (vectorized) AEC estimates (*brain* variables; 2556 unique connections). PLS estimates the maximal covariance between two different types of data blocks (Misic et al., 2016), which in our case would generate *saliences* representing the brain variables consisting of AEC estimates (brain saliences), and the experimental design variables describing the groups (design saliences). The output of PLS, referred to as latent variables (LV), is a composite of singular values, which describe the maximal covariance between the brain and the design variables, and two singular vectors with brain and design saliences. Saliences are the weights for each input data block. Linear combinations of brain saliences with the input data matrix provide brain scores per participants. For our group comparisons, two LVs per comparison were produced.

Statistical inference of LVs was determined by permutation tests with 1000 iterations with a threshold of p<0.05. These tests indicated which LVs were significant as an entire multivariate pattern. To identify the AECs making a significant contribution to the LV pattern, we employed bootstrapping with 500 resamples to compute the standard errors (SE) of brain saliences. Bootstrap ratios (BSR) were computed as a ratio of the brain saliences and the bootstrap SEs, which provided an estimate of the reliability of contributions of brain saliences. For significant LVs, BSR maps were interpreted at thresholds of ±3.5 or higher (Misic et al., 2016).

Next, behavioral PLS analyses were conducted within the stroke group for brain-behavior correlations between WAB subscale scores (fluency, comprehension, repetition, naming) and AEC estimates. Separate PLS analyses were conducted for correlations with each of the WAB measure. The methods for determining statistical significance and reliability of brain saliences were the same as the group PLS analysis procedures described above. The Rotman PLS toolbox in MATLAB (McIntosh et al., 1996) was used for both group and behavioral PLS analyses. To visualize the number of significant connections to nodes, a graph theory measure of node degree was computed (Figures 2 and 3).

**Figure 2.**
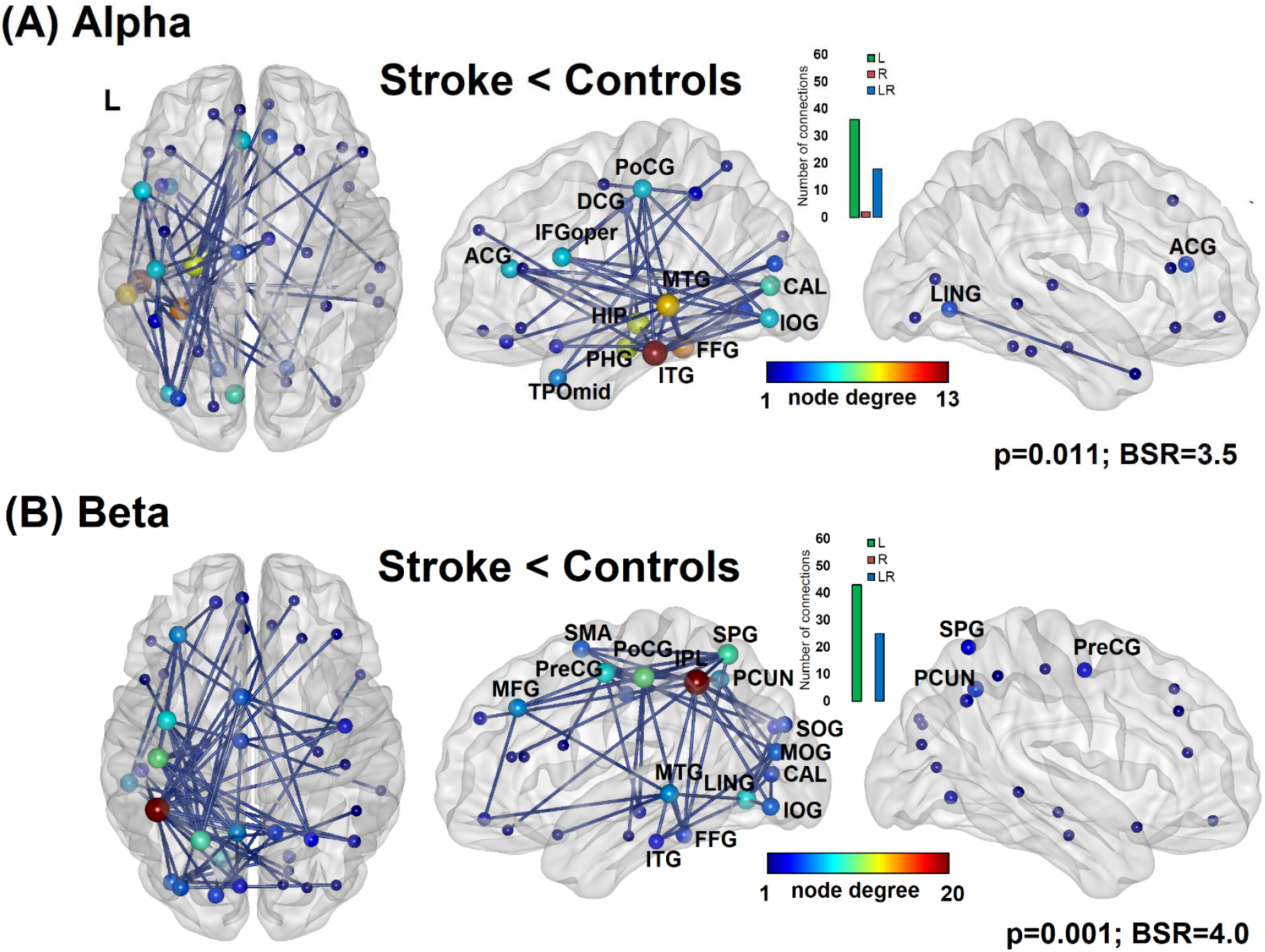
Spatial topology of *hypo*connectivity, as measured by amplitude envelope correlations, in the (A) alpha and (B) beta bands in stroke patients with aphasia compared to age-matched healthy controls. The size of the nodes indicates a graph theory measure of node degree. The inset displays the number of intrahemispheric left (L), right (R) or interhemispheric (LR) connections. The maps are displayed for bootstrap ratio (BSR) thresholds of 3.5 for alpha and 4.0 for beta connectivity.

**Figure 3.**
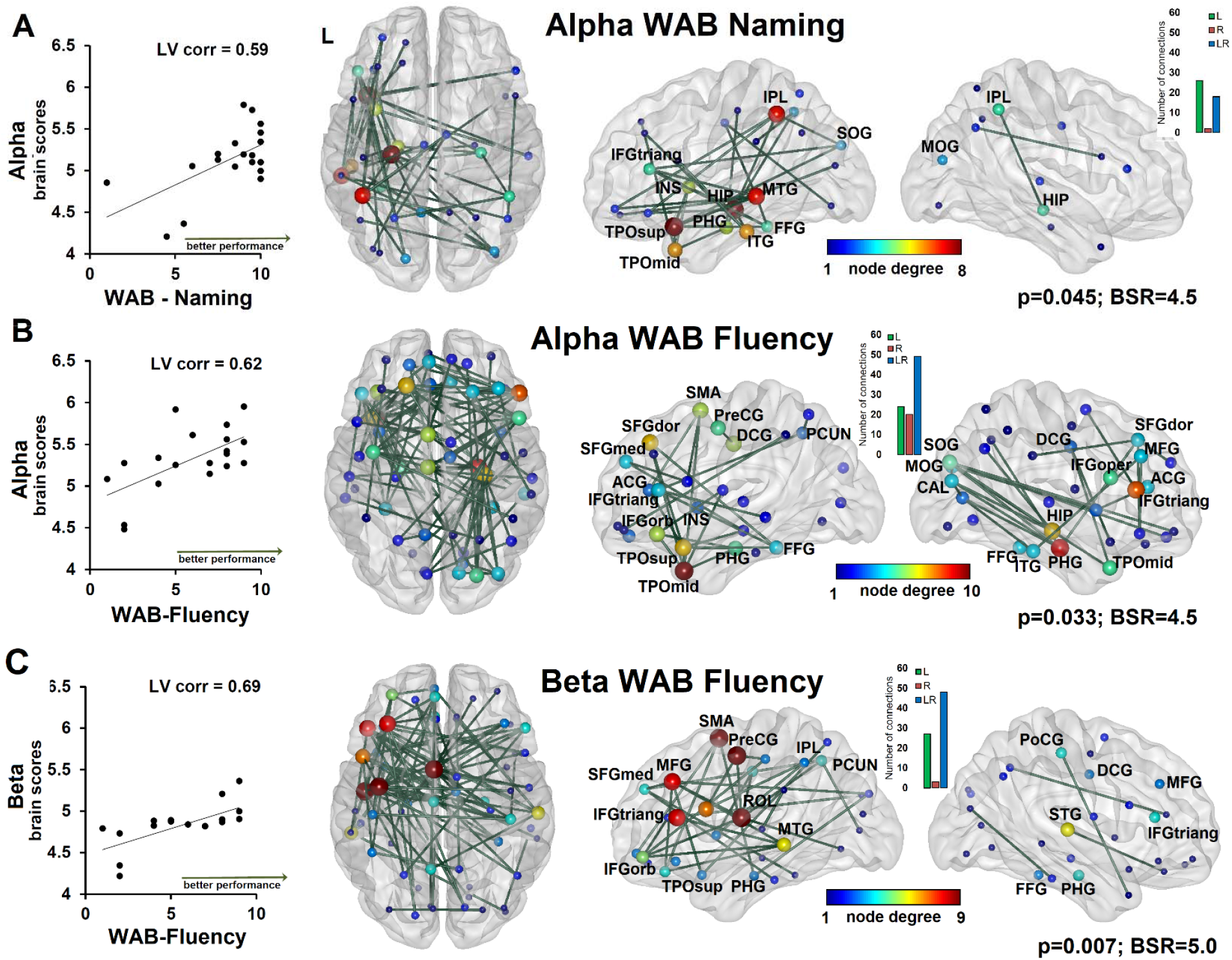
Spatial topology of positive correlations between alpha connectivity and (A) WAB object naming and (B) WAB fluency subscores, and between (C) beta connectivity and WAB fluency subscores in stroke survivors with aphasia. The size of the nodes indicates node degree. The inset displays the number of intrahemispheric left (L), right (R) or interhemispheric (LR) connections. The maps are displayed for bootstrap ratio (BSR) thresholds of 4.5 for alpha and 5.0 for beta connectivity.

Finally, a graph measure of node strength was computed based on statistically significant (or thresholded) connectivity matrices for both group differences and within-stroke correlations with WAB subscales. Brain Connectivity Toolbox (Rubinov and Sporns, 2010) was used to compute undirected node strengths by taking the sum of AEC values at each node. To evaluate how node strengths might be affected by local spectral properties (e.g., relative power) and by the extent of lesion within nodes, we ran node-wise, post-hoc regression analyses that were corrected for multiple comparisons using the False Discovery Rate (FDR) method (Benjamini and Hochberg, 1995). We controlled for relative power within a node for verifying group differences in node strengths by fitting a generalized linear model (GLM) with a binomial function; the group designation was the dependent variable and the relative power and node strength were the independent variables in this analysis [group ∼ relative power + node strength]. For correlations between node strengths and WAB subscores, we controlled for additional variables such as age, time post stroke onset, node lesion sizes along with relative power estimates in a linear regression analysis; the WAB subscore was the dependent variable and covariates and node strengths were the independent variables [wab_score ∼ age + time post onset + node lesion size + relative power + node strength]. RStudio was used for both types of posthoc analyses with *glm* (with family=binomial) or *lm* functions in the *stats* package.

## 3. RESULTS

The connectivity matrices and the spatial topology of connections representing the top 2-5% of connections in each of the frequency band of interest (theta, alpha, beta and low-gamma) are provided in Supplementary Figures 2 and 3 for healthy controls and stroke patients, respectively.

### 3.1 Group differences

PLS analysis revealed that the first LV was significant, indicating a significant effect of group for the AEC estimates in the alpha (p=0.011) and beta (p=0.001) frequency bands (Figure 2). None of the LVs was significant for theta or low-gamma AEC estimates (p>0.05). The direction of the differences pointed to hypoconnectivity in the alpha and beta bands in stroke patients compared to controls. As would be expected, the number of connections (estimated by AEC) on the side of stroke lesions was lower in patients. The spatial topology of reduced connectivity was distinct between alpha and beta bands, as can be visualized in Figure 2, with BSR thresholds of 3.5 and 4.0, respectively. Hypoconnectivity in the alpha band involved both left hemisphere ventral and dorsal nodes, including the inferior occipital (IOG), fusiform (FFG), middle and inferior temporal (MTG and ITG), parahippocampal (PHG) gyri, and the hippocampus (HIP), calcarine (CAL) cortex, temporal pole regions, the postcentral (PoCG), median (DCG) and anterior cingulate (ACG) and inferior frontal pars opercularis (IFGoper) gyri. The topology of beta hypoconnectivity involved primarily left dorsal nodes including the middle frontal (MFG), precentral and postcentral (PreCG and PoCG), superior parietal (SPG) gyri, supplementary motor areas (SMA), inferior parietal lobule (IPL), precuneus (PCUN), as well as the occcipital gyri, CAL, lingual gyrus (LING), and MTG, FFG and ITG. The posthoc regression analyses fitting a GLM revealed that differences in node strengths involving the left nodes remained significant after controlling for node relative power for both the alpha and beta bands; results are summarized in Supplementary Tables 1-2.

### 3.2 Correlations with WAB subscores

Alpha AEC or connectivity was positively correlated with object naming (p=0.045; LV correlation = 0.59, 95% CI: 0.54-0.83) and fluency (p=0.033; LV correlation = 0.62, 95% CI: 0.56-0.81) scores, and beta AEC was positively correlated with fluency scores (p=0.007; LV correlation=0.69, 95% CI: 0.67-0.87). PLS correlations with WAB repetition and auditory comprehension scores were not significant. A distinct topology of connections associated with significant correlations was found between alpha and beta bands (Figure 3). Stronger alpha connectivity among left MTG, FFG, ITG, PHG, IPL, IFG pars triangularis (IFGtriang), insula and temporal pole regions was correlated with better performance on WAB naming. Stronger alpha connectivity among bilateral dorsal and medial aspects of the superior frontal gyrus (SFGdor, SFGmed), left PreCG, bilateral DCG, PCUN, bilateral ACG, IFGtriang and IFG pars orbitalis (IFGorb), insula and temporal polar regions, along with PHG and FFG was associated with better performance on the WAB fluency measure. Finally, beta connectivity among left rolandic operculum (ROL), IFGtriang, IFGorb and IFGoper, middle frontal gyrus (MFG), PreCG, SMA, IPL, PCUN, MTG and PHG was associated with better performance on the WAB fluency measure.

Posthoc linear regression analyses indicated that node strengths derived from alpha and beta connectivity and WAB subscore relationships remained significant after controlling for age, time post stroke onset, node lesion sizes and node relative power estimates. The results from these analyses are summarized in Supplementary Tables 3-5. None of the PLS correlations between AEC and WAB subscores were significant in the theta and low-gamma bands.

## 4. DISCUSSION

In the current study, we quantified spontaneous electrophysiological connectivity in post-stroke chronic aphasia survivors using rsMEG with an overarching aim to examine reorganized connections between brain regions that critically support spared language functions after stroke. The methods included computations of amplitude envelope correlations or AEC, a commonly used measure of oscillatory coupling during resting-state (Brookes et al., 2011a; O’Neill et al., 2015), in frequency bands ranging from theta to low-gamma. The analyses of AECs revealed that the connectivity among (left) perilesional areas was greatly reduced in the alpha (8-13 Hz) and beta (15-30 Hz) bands in stroke survivors with aphasia compared to age-matched healthy controls. The spatial topology of hypoconnectivity between the alpha and beta bands was distinct, revealing a greater involvement of ventral frontal, temporal and parietal areas in alpha, and dorsal frontal and parietal areas in beta. In addition to the group differences, connectivity correlations with linguistic subscores from a clinical aphasia battery revealed the topology of connections that underlie preserved abilities to name pictures of objects and produce spontaneous speech after stroke. These results indicated that stronger alpha connectivity was associated with better naming performance, and stronger connectivity in both the alpha and beta bands was associated with better speech fluency performance (Figure 3). Consistent with the group results, the topology was distinct between these frequency bands, with alpha connections involving the IPL, MTG, ATL and ventral IFG regions as related to naming, whereas beta connections involving the PreCG, SMA, ROL and MFG as related to fluency. We controlled for lesion sizes by removing MEG signals within the lesioned tissue, assumed to be electrically-silent (Spironelli and Angrilli, 2009), in individual patients, before summarizing signals within parcels and prior to any statistical analysis. We also included nodal lesion sizes as a covariate in posthoc analyses with a graph theory measure of node strength. Overall, our results suggest a critical role of coherent activity within the alpha and beta bands, governing distinct language functions after stroke.

The involvement of alpha and beta that we found in the current study is consistent with multiple prior studies indicating an important role of spontaneous coherent activity in these frequency bands in post-stroke neuroplasticity (Dubovik et al., 2012; Westlake et al., 2012; Dubovik et al., 2013; Nicolo et al., 2015; Petrovic et al., 2017). To our knowledge, the findings related to the topology of alpha and beta hypoconnectivity and positive correlations with language performance are novel and as such contribute to our understanding of frequency-specific connectivity profiles in aphasia. These findings provide initial evidence of residual networks involving alpha and beta bands, underpinning linguistic abilities after stroke. One clinically-relevant future goal is to verify the repeatability of these patterns over multiple time points, and use this knowledge to advance our understanding of connectivity profiles within individuals and how they might change over the course of stroke recovery with and without targeted speech and language treatments.

Our use of MEG allowed computations of functional connectivity across multiple spatial and temporal scales. MEG is a powerful imaging modality (Baillet, 2017) that affords a direct evaluation of neuronal oscillatory activity—an “intrinsic physiological process” thought to mediate connectivity across brain regions and support cognitive processes (Engel et al., 2013; O’Neill et al., 2015). MEG facilitates computation of connectivity that is defined not only by the amplitude or envelope of a signal but also by its phase. In the current study, we quantified amplitude-based coupling or AEC to gain insights into the *resting-state* connectomes in post-stroke aphasia. Amplitude fluctuations are thought to capture slower aspects of interregional communication (Cox et al., 2018). Our focus on AEC was motivated by a number of prior studies that found a strong agreement between fMRI-derived resting-state networks (RSNs) and those derived from rsMEG using AEC (Brookes et al., 2011b; Hipp et al., 2012; Tewarie et al., 2014). In particular, AEC of alpha and beta oscillations (8-30 Hz) reveals patterns that are similar to fMRI RSNs (Cabral et al., 2014; Wei et al., 2021). The AEC estimates of connectivity are also found to be most consistent across repeated measurements both within participants and for group-level inferences compared to phase-based measures (Colclough et al., 2016). The physiological mechanisms that produce the temporally coordinated amplitude fluctuations measured by AEC remain unclear. Computational modeling studies provide some insight. For example, according to Cabral and colleagues (2014), the emergence of oscillatory envelope fluctuations is found to be linked to a “metastable synchronization regime” that is shaped by the precise space-time network structure of our brain. Temporary large-scale synchronization across different brain regions is possible in this regime at frequencies that typically fall within the alpha/beta range (Cabral et al., 2014). Full exploration of candidate mechanisms underlying AECs is one of the major themes of ongoing research in the field.

Dominant nodes depicted in the alpha connectivity correlations with naming as well as those in beta connectivity correlations with fluency scores are well-aligned with cognitive operations involving visual, language, motor/articulatory and executive control processes. The network exhibiting correlations with naming ability in alpha involved connections among the IPL, MTG, FFG, IFGtriang, ITG, temporal pole and medial temporal nodes, which are typically associated with visual recognition, semantic and phonological processing (Hickok and Poeppel, 2007; Binder et al., 2009; Price, 2012; Baldo et al., 2013; Binder, 2017). In contrast, the alpha-based fluency network involved bilateral superior and middle frontal gyri, anterior cingulate gyrus, SMA, precuneus and lateral IFG nodes that typically form the multiple-demand network, potentially subserving complex operations and sequential programming (Duncan, 2010) as required during the production of spontaneous and connected speech. The motor articulatory planning and execution processes during speech production appear to be captured in the beta connectivity correlations with fluency, which involve the rolandic operculum, SMA and precentral gyrus, and MFG, along with other frontal and parietal nodes (Knopman et al., 1983; Brown et al., 2005; Ackermann and Riecker, 2010; Biesbroek et al., 2016; Itabashi et al., 2016). The WAB battery that we used for our connectivity-behavior correlations is routinely used in clinical research studies for classification of aphasia subtypes but it does not isolate component language processes. While the nodes and connections as revealed from our analyses are consistent with prior fMRI and EEG/MEG, we acknowledge that the use of “multifactorial” measures such as WAB naming and fluency complicates our understanding of connectivity profiles underpinning constituent language processes. Naming is a complex multi-stage process that involves visual processing of the picture stimulus, visual recognition, access to meaning and phonological word forms, and motor planning and execution (DeLeon et al., 2007). Similarly, as described by Halai et al. (2017), it is unclear to what extent a measure of spontaneous speech by picture description reflects semantic or phonological skills (Halai et al., 2017). Therefore, a central future goal of our work to fully characterize frequency-specific connectivity/network properties is to identify unique correlates of constituent semantic and phonological processes in post-stroke aphasia. This knowledge would be critical for parsing out the effects of language treatments targeting one or more of these processes.

Complementary to amplitude coupling, as mentioned earlier, MEG connectivity can be depicted by examining synchronization of phases in source time series from different brain regions (Lachaux et al., 1999; Stam et al., 2007). While outside the scope of the current study, it would be equally interesting to evaluate phase synchronization differences in post-stroke aphasia, and the relationship with language outcomes after stroke. The patterns of phase coupling, while remain to be explored in future studies, could be reliably distinct from amplitude-based networks, thus revealing different aspects of neuronal dynamics (Cox et al., 2018) supporting language functions after stroke.

## 5. CONCLUSIONS

In this study, we systematically characterized large-scale oscillatory dynamics in post-stroke aphasia, comparing MEG connectivity patterns with healthy age-matched controls, with respect to language functions, and across a range of frequencies using an amplitude-based metric (or AEC). Perilesional left hemispheric regions exhibited hypoconnectivity in the alpha and beta bands. Importantly, unique connectivity profiles underpinning preserved linguistic abilities such as naming and fluency were identified, providing important insights into residual, frequency-specific language networks after stroke. Our future goals are to identify frequency-specific connectivity correlates of constituent semantic and phonological language processes, characterize how they change over the course of spontaneous and treatment-induced recovery, and finally apply phase-based connectivity metrics to more fully characterize oscillatory network dynamics in post-stroke aphasia. Electrophysiological connectivity in post-stroke aphasia remains largely unexplored, and while our current study addresses this important gap, more dedicated research is warranted in this field.

## Supporting information

all supplemental info

## Data Availability

All data produced in the present study are available upon reasonable request to the authors

## ACKNOWLEDGMENTS

This work was supported by the Catalyst grant (PI: Meltzer) and the trainee award (Shah-Basak) from the Canadian Partnership for Stroke Recovery, Heart and Stroke Foundation (Ottawa, ON Canada). We would like to extend our gratitude toward the patients, volunteers, and their families for participating in this study, and to the Aphasia Institute and the March of Dimes York-Durham Aphasia Centre.

## SUPPLEMENTARY MATERIAL

**Supplementary Figure 1**. Seventy-two cortical and subcortical regions, as defined by the Automated Anatomical Labeling (AAL) atlas (Tzourio-Mazoyer et al., 2002) for AEC analyses.

**Supplementary Figures 2 and 3**. Connectivity matrices and spatial topology of connections representing the top 2-5% of connections in each of the frequency band of interest (theta, alpha, beta and low-gamma) in healthy controls (Supp. Fig. 2) and stroke survivors with aphasia (Supp. Fig. 3).

**Supplementary Table 1**. Estimated parameters from the binomial GLM analysis of node strengths of significant group differences in **<u>alpha</u>** connectivity, after controlling for nodewise relative power. Mean node strengths by group and FDR-corrected p-values are also provided.

**Supplementary Table 2**. Estimated parameters from the binomial GLM analysis of node strengths of significant group differences in **<u>beta</u>** connectivity, after controlling for nodewise relative power. Mean node strengths by group and FDR-corrected p-values are also provided.

**Supplementary Table 3**. Estimated parameters from the linear regression analysis of node strengths from significant **<u>alpha</u>** connectivity correlations with **WAB naming**, after controlling for age, time post stroke onset, nodewise relative power and lesion size. Mean node strengths in the stroke group, and FDR-corrected p-values are also provided.

**Supplementary Table 4**. Estimated parameters from the linear regression analysis of node strengths from significant **<u>alpha</u>** connectivity correlations with **WAB fluency**, after controlling for age, time post stroke onset, nodewise relative power and lesion size. Mean node strengths in the stroke group, and FDR-corrected p-values are also provided.

**Supplementary Table 5**. Estimated parameters from the linear regression analysis of node strengths from significant **<u>beta</u>** connectivity correlations with **WAB fluency**, after controlling for age, time post stroke onset, nodewise relative power and lesion size. Mean node strengths in the stroke group, and FDR-corrected p-values are also provided.

## Notes

### Competing Interest Statement

The authors have declared no competing interest.

### Funding Statement

This study was funded by the Catalyst award (PI: Meltzer) and a trainee award (PI: Shah-Basak) from the Canadian Partnership for Stroke Recovery, Heart and Stroke Foundation, Ottawa, ON CANADA

### Author Declarations

Research Ethics Board of Rotman Research Institute at Baycrest Health Sciences gave ethical approval for this work.

