## Supplementary material for "Electrophysiological connectivity markers of preserved language functions in post-stroke aphasia": all supplemental info

**Supplementary Figure 1.** Seventy-two cortical and subcortical regions, as defined by the Automated Anatomical Labeling (AAL) atlas (Tzourio-Mazoyer et al., 2002) for AEC analyses.

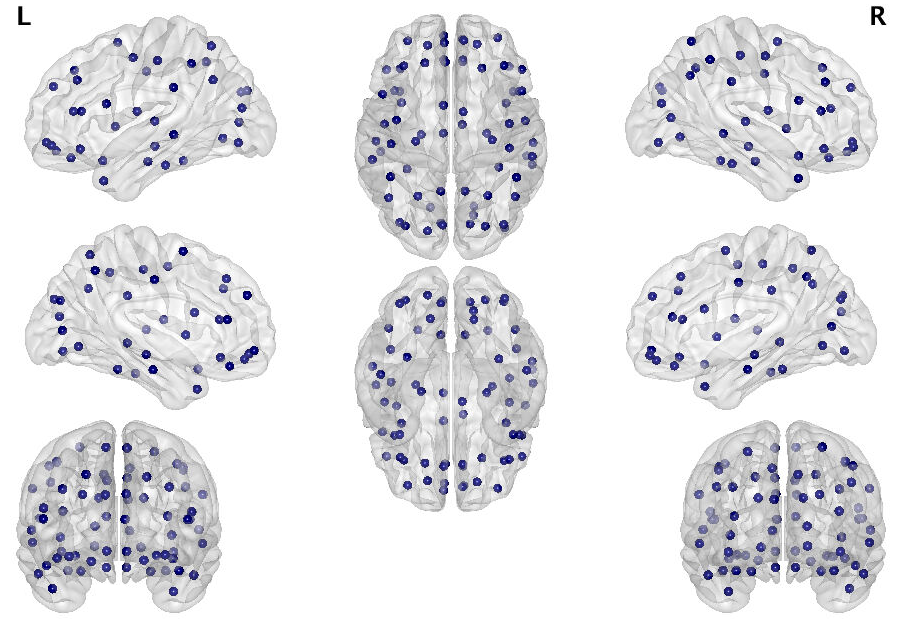

|  | **Abbreviations** | **Regions** |  | **Abbreviations** | **Regions** |
| --- | --- | --- | --- | --- | --- |
|  | L | Left | 18 | PHG | Parahippocampal gyrus |
|  | R | Right | 19 | CAL | Calcarine |
| 1 | PreCG | Precentral gyrus | 20 | CUN | Cuneus |
| 2 | SFGdor | Superior frontal gyrus, dorsolateral | 21 | LING | Lingual gyrus |
| 3 | ORBsup | Orbital gyrus, superior | 22 | SOG | Superior occipital gyrus |
| 4 | MFG | Middle frontal gyrus | 23 | MOG | Middle occipital gyrus |
| 5 | ORBmid | Orbital gyrus, middle | 24 | IOG | Inferior occipital gyrus |
| 6 | IFGoperc | Inferior frontal gyrus, pars opercularis | 25 | FFG | Fusiform gyrus |
| 7 | IFGtriang | Inferior frontal gyrus, pars triangularis | 26 | PoCG | Postcentral gyrus |
| 8 | ORBinf | Inferior frontal gyrus, pars orbitalis | 27 | SPG | Superior parietal lobe |
| 9 | ROL | Rolandic operculum | 28 | IPL | Inferior parietal lobe |
| 10 | SMA | Supplementary motor area | 29 | SMG | Supramarginal gyrus |
| 11 | SFGmed | Superior frontal gyrus, medial | 30 | ANG | Angular gyrus |
| 12 | ORBsupmed | Orbital gyrus, superior medial | 31 | PCUN | Precuneus |
| 13 | REC | Rectus gyrus | 32 | STG | Superior temporal gyrus |
| 14 | INS | Insula | 33 | TPOsup | Temporal pole, superior |
| 15 | ACG | Anterior cingulate gyrus | 34 | MTG | Middle temporal gyrus |
| 16 | DCG | Middle cingulate gyrus | 35 | TPOmid | Temporal pole, middle |
| 17 | HIP | Hippocampus | 36 | ITG | Inferior temporal gyrus |

**Supplementary Figures 2 and 3.** Connectivity matrices and spatial topology of connections representing the top 2-5% of connections in each of the frequency band of interest (theta, alpha, beta and low-gamma) in healthy controls (Supp. Fig. 2) and stroke survivors with aphasia (Supp. Fig. 3).

**Supp. Fig. 2**

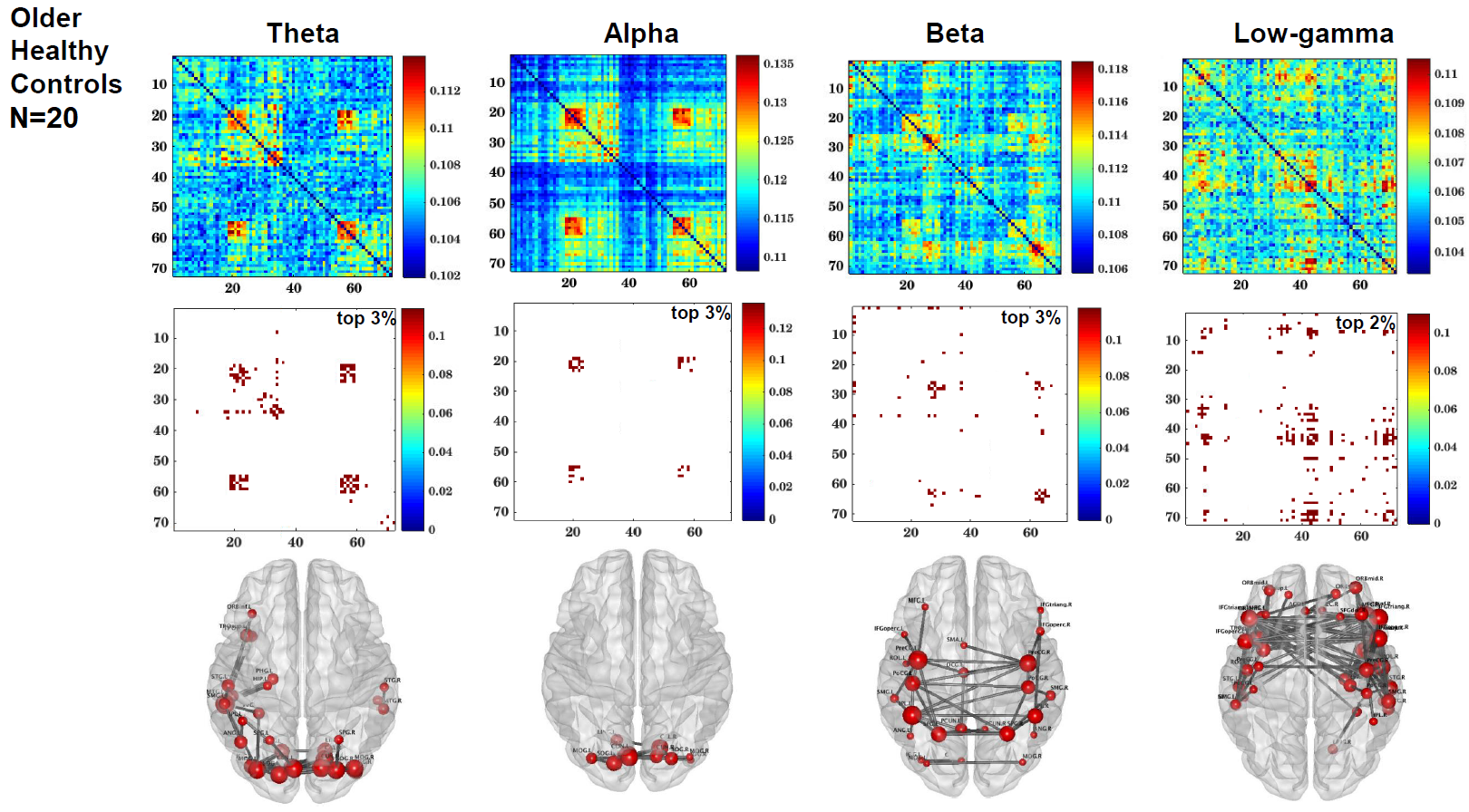

**Supp. Fig. 3**

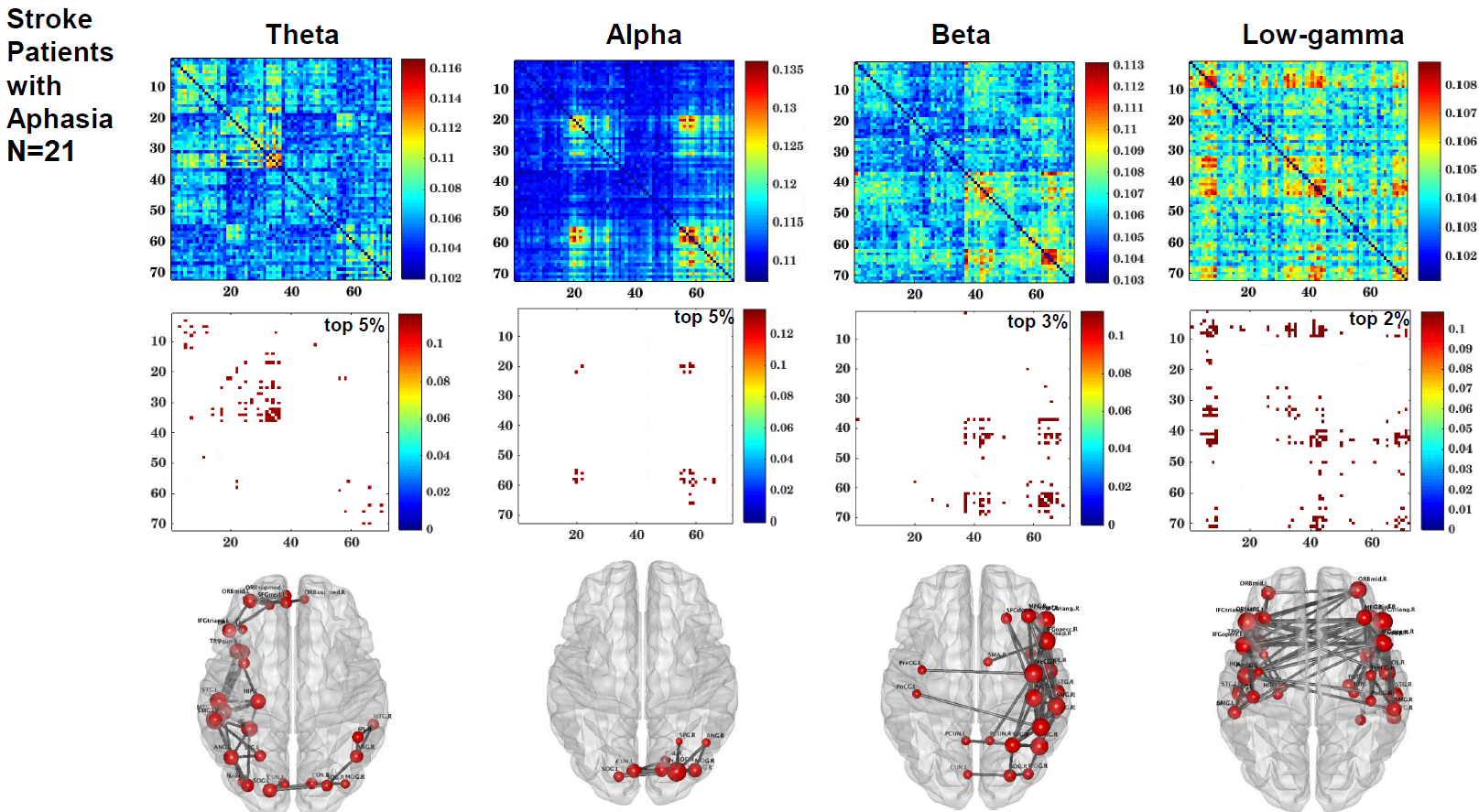

**Software packages used in this study:**

Partial least squares analyses: <https://www.rotman-baycrest.on.ca/index.php?section=84>

MNE-Python package: <https://mne.tools/stable/index.html>

BrainNet viewer: <https://github.com/mingruixia/BrainNet-Viewer>

Brain Connectivity Toolbox: <https://sites.google.com/site/bctnet/>

**Supplementary Table 1.** Estimated parameters from the binomial GLM analysis of node strengths of significant group differences in **alpha** connectivity, after controlling for nodewise relative power. Mean node strengths by group and FDR-corrected p-values are also provided.

| **AAL node** | **Mean**  **node strength (stroke)** | **Mean**  **node strength (controls)** | **Estimate** | **z value** | **FDR p-value** |
| --- | --- | --- | --- | --- | --- |
| ITG.L | 1.49 | 1.64 | -11.03 | -3.13 | 0.011 |
| PHG.L | 1.02 | 1.12 | -23.54 | -3.04 | 0.011 |
| MTG.L | 0.80 | 0.89 | -19.75 | -2.95 | 0.011 |
| FFG.L | 0.91 | 1.00 | -20.88 | -2.60 | 0.013 |
| HIP.L | 0.67 | 0.73 | -29.16 | -2.84 | 0.011 |
| CAL.L | 0.69 | 0.74 | -23.50 | -2.11 | 0.035 |
| TPOsup.L | 0.45 | 0.49 | -38.78 | -2.80 | 0.011 |
| TPOmid.L | 0.44 | 0.48 | -59.44 | -2.89 | 0.011 |
| ACG.L | 0.44 | 0.48 | -47.89 | -2.99 | 0.011 |
| MOG.L | 0.34 | 0.37 | -47.16 | -2.96 | 0.011 |
| LING.R | 0.34 | 0.37 | -41.41 | -2.69 | 0.011 |
| ACG.R | 0.33 | 0.36 | -62.67 | -2.80 | 0.011 |
| LING.L | 0.23 | 0.26 | -79.33 | -3.00 | 0.011 |
| IPL.L | 0.22 | 0.24 | -72.50 | -3.03 | 0.011 |
| PoCG.L | 0.22 | 0.24 | -55.29 | -2.16 | 0.032 |
| REC.L | 0.22 | 0.24 | -60.05 | -2.46 | 0.016 |
| IFGoperc.L | 0.22 | 0.24 | -104.07 | -2.94 | 0.011 |
| PHG.R | 0.22 | 0.24 | -74.44 | -2.77 | 0.011 |
| ORBinf.R | 0.22 | 0.24 | -82.23 | -2.81 | 0.011 |
| STG.L | 0.12 | 0.13 | -99.61 | -2.75 | 0.011 |
| INS.L | 0.11 | 0.13 | -120.45 | -2.68 | 0.011 |
| SOG.L | 0.11 | 0.13 | -134.48 | -2.96 | 0.011 |
| PreCG.L | 0.11 | 0.12 | -110.48 | -2.41 | 0.017 |
| IOG.L | 0.12 | 0.13 | -104.58 | -2.53 | 0.014 |
| MTG.R | 0.11 | 0.12 | -118.17 | -2.83 | 0.011 |
| DCG.L | 0.11 | 0.12 | -116.24 | -2.87 | 0.011 |
| IFGtriang.L | 0.11 | 0.12 | -116.43 | -2.68 | 0.011 |
| ITG.R | 0.12 | 0.13 | -99.58 | -2.73 | 0.011 |
| DCG.R | 0.11 | 0.12 | -107.09 | -2.55 | 0.014 |
| IOG.R | 0.11 | 0.12 | -166.54 | -2.85 | 0.011 |
| HIP.R | 0.11 | 0.12 | -96.19 | -2.36 | 0.019 |
| IFGtriang.R | 0.11 | 0.12 | -184.21 | -2.42 | 0.017 |
| SPG.L | 0.11 | 0.12 | -108.06 | -2.58 | 0.014 |
| FFG.R | 0.11 | 0.12 | -223.25 | -2.57 | 0.014 |
| ORBsupmed.R | 0.11 | 0.12 | -153.01 | -2.70 | 0.011 |
| CAL.R | 0.12 | 0.12 | -98.84 | -2.52 | 0.014 |
| ORBinf.L | 0.11 | 0.12 | -176.72 | -2.82 | 0.011 |
| SFGmed.L | 0.11 | 0.12 | -184.69 | -2.66 | 0.012 |
| TPOmid.R | 0.11 | 0.12 | -119.41 | -2.49 | 0.015 |

**Supplementary Table 2**. Estimated parameters from the binomial GLM analysis of node strengths of significant group differences in **beta** connectivity, after controlling for nodewise relative power. Mean node strengths by group and FDR-corrected p-values are also provided.

| **AAL node** | **Mean**  **node strength (stroke)** | **Mean**  **node strength**  **(controls)** | **Estimate** | **z value** | **FDR p-value** |
| --- | --- | --- | --- | --- | --- |
| IPL.L | 2.02 | 2.18 | -22.22 | -3.08 | 0.008 |
| SPG.L | 0.95 | 1.04 | -55.57 | -3.06 | 0.008 |
| PreCG.L | 0.96 | 1.04 | -36.49 | -2.90 | 0.008 |
| PoCG.L | 0.96 | 1.04 | -52.18 | -2.31 | 0.026 |
| IOG.L | 0.63 | 0.68 | -60.91 | -2.79 | 0.010 |
| CAL.L | 0.64 | 0.68 | -61.38 | -2.85 | 0.009 |
| PCUN.L | 0.53 | 0.58 | -76.65 | -3.10 | 0.008 |
| MOG.L | 0.52 | 0.57 | -83.00 | -2.81 | 0.010 |
| LING.L | 0.53 | 0.57 | -49.27 | -2.44 | 0.020 |
| DCG.L | 0.43 | 0.47 | -96.92 | -2.33 | 0.025 |
| SMA.L | 0.42 | 0.46 | -89.84 | -3.05 | 0.008 |
| PCUN.R | 0.32 | 0.35 | -78.48 | -2.90 | 0.008 |
| SOG.L | 0.32 | 0.34 | -115.23 | -3.01 | 0.008 |
| ANG.R | 0.32 | 0.34 | -158.84 | -1.98 | **0.053** |
| SPG.R | 0.32 | 0.34 | -104.57 | -2.22 | 0.031 |
| MFG.L | 0.32 | 0.34 | -128.57 | -3.01 | 0.008 |
| MTG.L | 0.31 | 0.34 | -134.13 | -2.93 | 0.008 |
| FFG.L | 0.31 | 0.34 | -139.04 | -2.76 | 0.010 |
| PreCG.R | 0.21 | 0.23 | -130.53 | -2.56 | 0.015 |
| HIP.L | 0.21 | 0.23 | -338.27 | -2.99 | 0.008 |
| LING.R | 0.21 | 0.23 | -122.01 | -2.00 | **0.052** |
| ITG.L | 0.21 | 0.23 | -212.08 | -2.63 | 0.013 |
| SFGmed.L | 0.21 | 0.23 | -235.10 | -3.06 | 0.008 |
| CUN.L | 0.21 | 0.23 | -220.15 | -2.72 | 0.011 |
| ACG.L | 0.21 | 0.23 | -186.29 | -1.96 | **0.054** |
| ORBsup.L | 0.21 | 0.22 | -216.12 | -2.90 | 0.008 |
| PHG.L | 0.21 | 0.22 | -196.08 | -2.85 | 0.009 |
| CAL.R | 0.11 | 0.11 | -282.20 | -2.72 | 0.011 |
| PoCG.R | 0.11 | 0.12 | -214.45 | -3.12 | 0.008 |
| MTG.R | 0.11 | 0.11 | -321.15 | -3.17 | 0.008 |
| IFGoperc.L | 0.11 | 0.12 | -183.97 | -2.99 | 0.008 |
| IPL.R | 0.10 | 0.11 | -210.05 | -1.72 | **0.088** |
| MOG.R | 0.11 | 0.12 | -196.31 | -2.92 | 0.008 |
| SFGdor.R | 0.11 | 0.11 | -185.89 | -2.96 | 0.008 |
| REC.L | 0.10 | 0.11 | -277.05 | -2.39 | 0.022 |
| SOG.R | 0.11 | 0.11 | -248.07 | -2.43 | 0.020 |
| ORBsup.R | 0.10 | 0.11 | -283.03 | -2.30 | 0.026 |
| CUN.R | 0.11 | 0.11 | -228.89 | -2.97 | 0.008 |
| ACG.R | 0.11 | 0.11 | -247.53 | -2.70 | 0.011 |
| MFG.R | 0.11 | 0.11 | -189.90 | -1.51 | **0.132** |
| TPOsup.L | 0.10 | 0.11 | -204.79 | -1.91 | **0.059** |

**Supplementary Table 3**. Estimated parameters from the linear regression analysis of node strengths from significant **alpha** connectivity correlations with **WAB** **naming,** after controlling for age, time post stroke onset, nodewise relative power and lesion size. Mean node strengths in the stroke group, and FDR-corrected p-values are also provided.

| **AAL node** | **Mean node strength  (stroke)** | **Estimate** | **t value** | **FDR p-value** |
| --- | --- | --- | --- | --- |
| TPOsup.L | 0.79 | 32.49 | 4.46 | 0.006 |
| MTG.L | 0.69 | 32.39 | 3.46 | 0.014 |
| HIP.L | 0.69 | 22.13 | 3.32 | 0.014 |
| ITG.L | 0.68 | 32.62 | 3.01 | 0.017 |
| IPL.R | 0.59 | 29.63 | 3.09 | 0.016 |
| IPL.L | 0.57 | 41.85 | 3.40 | 0.014 |
| HIP.R | 0.46 | 57.24 | 4.82 | 0.003 |
| FFG.L | 0.46 | 46.73 | 4.05 | 0.010 |
| TPOmid.L | 0.45 | 48.59 | 3.83 | 0.012 |
| SOG.L | 0.34 | 57.21 | 2.71 | 0.025 |
| INS.L | 0.34 | 47.14 | 2.90 | 0.018 |
| PHG.L | 0.34 | 63.74 | 3.30 | 0.014 |
| SPG.L | 0.23 | 52.28 | 2.59 | 0.031 |
| PCUN.L | 0.23 | 71.81 | 3.22 | 0.016 |
| MOG.R | 0.23 | 33.67 | 3.14 | 0.016 |
| ANG.R | 0.23 | 80.35 | 3.65 | 0.012 |
| ORBinf.L | 0.23 | 44.15 | 1.86 | **0.087** |
| PHG.R | 0.23 | 45.08 | 1.84 | **0.087** |
| ORBmid.L | 0.23 | 76.66 | 3.53 | 0.014 |
| DCG.R | 0.23 | 59.89 | 2.55 | 0.032 |
| IFGtriang.L | 0.23 | 66.44 | 2.48 | 0.034 |
| DCG.L | 0.23 | 97.71 | 3.53 | 0.014 |
| IFGtriang.R | 0.23 | 105.31 | 3.66 | 0.012 |
| ITG.R | 0.12 | 96.94 | 2.02 | **0.067** |
| ANG.L | 0.12 | 70.46 | 2.48 | 0.034 |
| ORBsup.L | 0.11 | 112.18 | 3.07 | 0.017 |
| IFGoperc.R | 0.11 | 169.64 | 5.26 | 0.002 |
| MOG.L | 0.11 | 206.45 | 3.30 | 0.014 |
| SPG.R | 0.11 | 169.43 | 2.45 | 0.034 |
| IOG.R | 0.11 | 166.39 | 2.06 | **0.063** |
| IOG.L | 0.11 | 170.03 | 2.95 | 0.018 |
| STG.R | 0.11 | 210.77 | 3.06 | 0.016 |
| MFG.L | 0.11 | 142.08 | 2.21 | **0.051** |
| TPOmid.R | 0.11 | 155.36 | 2.98 | 0.017 |
| IFGoperc.L | 0.11 | 107.90 | 2.90 | 0.018 |
| PoCG.L | 0.11 | 147.39 | 2.36 | 0.040 |
| SFGdor.L | 0.11 | 227.61 | 7.00 | 0.000 |

**Supplementary Table 4**. Estimated parameters from the linear regression analysis of node strengths from significant **alpha** connectivity correlations with **WAB** **fluency,** after controlling for age, time post stroke onset, nodewise relative power and lesion size. Mean node strengths in the stroke group, and FDR-corrected p-values are also provided.

| **AAL node** | **Mean node strength  (stroke)** | **Estimate** | **t value** | **FDR p-value** |
| --- | --- | --- | --- | --- |
| TPOsup.L | 1.01 | 34.15 | 5.54 | 0.001 |
| HIP.R | 0.94 | 32.27 | 4.83 | 0.002 |
| PHG.R | 0.92 | 31.72 | 3.74 | 0.006 |
| DCG.L | 0.90 | 37.32 | 4.37 | 0.003 |
| IFGtriang.R | 0.89 | 36.42 | 4.26 | 0.003 |
| PreCG.L | 0.89 | 34.40 | 3.49 | 0.008 |
| TPOmid.L | 0.88 | 25.16 | 5.10 | 0.002 |
| SMA.L | 0.78 | 40.37 | 3.68 | 0.007 |
| SFGdor.L | 0.77 | 52.93 | 5.56 | 0.001 |
| FFG.R | 0.68 | 31.72 | 3.98 | 0.005 |
| TPOmid.R | 0.57 | 55.41 | 4.80 | 0.002 |
| ORBinf.L | 0.56 | 38.83 | 2.71 | 0.023 |
| IFGoperc.R | 0.56 | 45.40 | 3.84 | 0.005 |
| DCG.R | 0.56 | 70.32 | 5.71 | 0.001 |
| CUN.R | 0.48 | 42.45 | 2.77 | 0.020 |
| SOG.R | 0.47 | 61.14 | 4.27 | 0.003 |
| PCUN.L | 0.47 | 43.48 | 3.31 | 0.009 |
| ITG.R | 0.46 | 53.04 | 3.90 | 0.005 |
| PHG.L | 0.46 | 61.66 | 4.76 | 0.002 |
| MFG.R | 0.45 | 65.63 | 3.59 | 0.007 |
| IFGtriang.L | 0.45 | 43.53 | 2.51 | 0.029 |
| FFG.L | 0.45 | 65.52 | 4.26 | 0.003 |
| SFGmed.L | 0.45 | 80.99 | 3.99 | 0.005 |
| ACG.R | 0.45 | 107.37 | 5.50 | 0.001 |
| INS.L | 0.45 | 55.20 | 3.50 | 0.008 |
| MOG.R | 0.39 | 57.11 | 3.83 | 0.005 |
| CAL.R | 0.37 | 62.84 | 3.77 | 0.006 |
| SPG.L | 0.35 | 54.59 | 3.56 | 0.008 |
| MTG.L | 0.34 | 110.31 | 6.25 | 0.001 |
| INS.R | 0.34 | 76.77 | 2.96 | 0.016 |
| ORBsup.L | 0.34 | 62.02 | 2.65 | 0.025 |
| SFGdor.R | 0.33 | 81.69 | 3.30 | 0.009 |
| ACG.L | 0.33 | 101.10 | 4.56 | 0.003 |
| IFGoperc.L | 0.33 | 54.75 | 2.62 | 0.025 |
| CAL.L | 0.23 | 117.69 | 3.36 | 0.009 |
| STG.R | 0.23 | 147.56 | 4.62 | 0.002 |
| ORBmid.L | 0.23 | 121.19 | 4.78 | 0.002 |
| LING.R | 0.23 | 133.64 | 3.39 | 0.009 |
| ORBsup.R | 0.22 | 132.48 | 5.24 | 0.001 |
| HIP.L | 0.22 | 94.12 | 2.83 | 0.019 |
| REC.R | 0.22 | 114.90 | 3.41 | 0.009 |
| MFG.L | 0.22 | 100.53 | 2.24 | 0.045 |
| ORBsupmed.L | 0.22 | 116.51 | 3.18 | 0.010 |
| ORBinf.R | 0.22 | 80.44 | 2.06 | **0.059** |
| PreCG.R | 0.22 | 152.32 | 3.33 | 0.009 |
| SFGmed.R | 0.22 | 131.93 | 3.13 | 0.011 |
| SMA.R | 0.22 | 135.63 | 3.40 | 0.009 |
| ANG.R | 0.12 | 205.66 | 3.73 | 0.006 |
| CUN.L | 0.12 | 141.28 | 3.21 | 0.010 |
| PCUN.R | 0.12 | 201.90 | 2.67 | 0.024 |
| SPG.R | 0.12 | 195.27 | 3.27 | 0.009 |
| STG.L | 0.12 | 134.69 | 2.96 | 0.016 |
| ROL.R | 0.11 | 166.97 | 2.49 | 0.029 |
| LING.L | 0.11 | 187.43 | 2.56 | 0.027 |
| ORBmid.R | 0.11 | 151.14 | 2.43 | 0.032 |
| ROL.L | 0.11 | 130.97 | 2.69 | 0.025 |
| IOG.R | 0.11 | 183.32 | 2.11 | **0.054** |
| IPL.R | 0.11 | 199.32 | 2.28 | 0.041 |
| TPOsup.R | 0.11 | 184.65 | 3.17 | 0.010 |
| ITG.L | 0.11 | 239.59 | 2.93 | 0.016 |
| ORBsupmed.R | 0.11 | 136.39 | 2.17 | 0.049 |
| IPL.L | 0.11 | 212.89 | 3.37 | 0.009 |
| PoCG.R | 0.11 | 187.41 | 2.61 | 0.025 |

**Supplementary Table 5**. Estimated parameters from the linear regression analysis of node strengths from significant **beta** connectivity correlations with **WAB** **fluency,** after controlling for age, time post stroke onset, nodewise relative power and lesion size. Mean node strengths in the stroke group, and FDR-corrected p-values are also provided.

| **AAL node** | **Mean**  **node strength  (stroke)** | **Estimate** | **t value** | **FDR p-value** |
| --- | --- | --- | --- | --- |
| ROL.L | 0.97 | 40.46 | 3.74 | 0.004 |
| IFGoperc.L | 0.86 | 46.62 | 4.01 | 0.003 |
| SMA.L | 0.86 | 55.34 | 4.37 | 0.002 |
| IFGtriang.L | 0.85 | 43.20 | 2.21 | 0.046 |
| MFG.L | 0.84 | 34.37 | 4.46 | 0.002 |
| PreCG.L | 0.75 | 39.87 | 5.09 | 0.001 |
| ORBmid.L | 0.73 | 36.44 | 5.01 | 0.001 |
| STG.R | 0.64 | 98.61 | 6.83 | 0.000 |
| MTG.L | 0.52 | 108.78 | 6.08 | 0.000 |
| PoCG.R | 0.43 | 125.83 | 6.00 | 0.000 |
| SFGmed.L | 0.43 | 115.02 | 3.84 | 0.003 |
| REC.L | 0.43 | 136.45 | 4.99 | 0.001 |
| PCUN.L | 0.42 | 42.97 | 4.12 | 0.002 |
| FFG.R | 0.41 | 51.19 | 5.62 | 0.001 |
| PHG.R | 0.41 | 45.84 | 4.15 | 0.002 |
| PreCG.R | 0.33 | 105.73 | 3.49 | 0.005 |
| ORBsupmed.L | 0.32 | 164.00 | 4.87 | 0.001 |
| INS.L | 0.32 | 110.33 | 2.64 | 0.023 |
| IFGtriang.R | 0.32 | 180.02 | 5.05 | 0.001 |
| TPOsup.L | 0.32 | 196.42 | 6.80 | 0.000 |
| IPL.L | 0.32 | 156.16 | 5.30 | 0.001 |
| ORBinf.L | 0.32 | 162.86 | 3.81 | 0.003 |
| MFG.R | 0.32 | 46.89 | 4.05 | 0.002 |
| PHG.L | 0.32 | 164.03 | 4.48 | 0.002 |
| ANG.R | 0.32 | 171.26 | 6.08 | 0.000 |
| CUN.R | 0.32 | 145.52 | 3.50 | 0.005 |
| DCG.L | 0.31 | 47.11 | 3.98 | 0.003 |
| IFGoperc.R | 0.22 | 177.22 | 4.54 | 0.001 |
| SMA.R | 0.22 | 207.88 | 3.88 | 0.003 |
| PoCG.L | 0.21 | 154.59 | 3.05 | 0.011 |
| ACG.L | 0.21 | 226.76 | 4.85 | 0.001 |
| ITG.R | 0.21 | 273.86 | 9.17 | 0.000 |
| DCG.R | 0.21 | 173.27 | 3.88 | 0.003 |
| HIP.R | 0.21 | 319.76 | 5.02 | 0.001 |
| PCUN.R | 0.21 | 246.34 | 4.67 | 0.001 |
| ITG.L | 0.21 | 193.29 | 2.75 | 0.019 |
| SOG.L | 0.21 | 222.39 | 4.25 | 0.002 |
| SPG.L | 0.21 | 246.74 | 4.03 | 0.003 |
| ORBsup.L | 0.21 | 229.65 | 4.37 | 0.002 |
| MOG.R | 0.11 | 367.58 | 4.84 | 0.001 |
| MTG.R | 0.11 | 357.96 | 3.40 | 0.005 |
| ROL.R | 0.11 | 308.96 | 4.03 | 0.002 |
| ORBmid.R | 0.11 | 241.94 | 2.82 | 0.016 |
| SFGdor.L | 0.11 | 360.35 | 2.57 | 0.025 |
| REC.R | 0.11 | 408.65 | 4.30 | 0.002 |
| MOG.L | 0.11 | 302.95 | 3.29 | 0.007 |
| SOG.R | 0.11 | 371.42 | 4.63 | 0.001 |
| ORBsup.R | 0.11 | 240.69 | 2.42 | 0.032 |
| ACG.R | 0.11 | 367.34 | 4.08 | 0.002 |
| FFG.L | 0.11 | 249.28 | 2.41 | 0.033 |
| HIP.L | 0.11 | 466.52 | 3.46 | 0.005 |
| CAL.R | 0.11 | 486.33 | 3.67 | 0.004 |
| TPOsup.R | 0.11 | 381.18 | 4.30 | 0.002 |
| LING.R | 0.11 | 500.05 | 4.22 | 0.002 |
| CAL.L | 0.11 | 332.21 | 3.80 | 0.003 |
| LING.L | 0.11 | 230.45 | 2.37 | 0.039 |
| CUN.L | 0.11 | 384.31 | 3.66 | 0.004 |
| STG.L | 0.11 | 252.16 | 2.36 | 0.039 |
| SMG.L | 0.11 | 346.21 | 3.96 | 0.003 |
| INS.R | 0.11 | 423.12 | 3.98 | 0.003 |
| IOG.R | 0.10 | 314.35 | 3.20 | 0.008 |
